# Stable Sparse Classifiers predict cognitive impairment from gait patterns

**DOI:** 10.1101/2022.03.10.22272227

**Authors:** Tania Aznielle-Rodríguez, Marlis Ontivero-Ortega, Lídice Galán-García, Hichem Sahli, Mitchell Valdés-Sosa

**Affiliations:** Department of Electronics, Cuban Center for Neuroscience, Havana, Cuba; Electronics and Informatics Department, Vrije Universiteit Brussels, Brussels, Belgium; Department of Neuroinformatics, Cuban Center for Neuroscience, Havana, Cuba; Department of Data Analysis, Faculty of Psychological and Educational Sciences, Ghent University, Ghent, Belgium; Interuniversity Microelectronics Centre, Heverlee, Belgium; Department of Cognitive Neuroscience, Cuban Center for Neuroscience, Havana, Cuba

**Author notes:** These authors have contributed equally to this work and share senior’s authorship (Hichem Sahli, Mitchell Valdés-Sosa).

**Keywords:** Gait analysis, cognitive impairment, classification, glmnet, stability ROC, biomarkers

## Abstract

**Background:** Although gait patterns disturbances are known to be related to cognitive decline, there is no consensus on the possibility of predicting one from the other. It is necessary to find the optimal gait features, experimental protocols, and computational algorithms to achieve this purpose.

**Purposes:** To assess the efficacy of the Stable Sparse Classifiers procedure (SSC) for discriminating young and older adults, as well as healthy and cognitively impaired elderly groups from their gait patterns. To identify the walking tasks or combinations of tasks and specific spatio-temporal gait features (STGF) that allow the best prediction with SSC.

**Methods:** A sample of 125 participants (40 young- and 85 older-adults) was studied. They underwent assessment with five neuropsychological tests that explore different cognitive domains. A summarized cognitive index (MDCog), based on the Mahalanobis distance from normative data, was calculated. The sample was divided into three groups (young adults, healthy and cognitively impaired elderly adults) using k-means clustering of MDCog in addition to Age. The participants executed four walking tasks (normal, fast, easy- and hard-dual tasks) and their gait patterns, measured with a body-fixed Inertial Measurement Unit, were used to calculate 16 STGF and dual-task costs. SSC was then employed to predict which group the participants belonged to. The classification’s performance was assessed using the area under the receiver operating curves (AUC). The set of STGF features and tasks producing the most accurate classifications were identified.

**Results:** The comparison between the three groups revealed significant differences for all STGF in all tasks, while the global AUC of the classification using SSC was 0.87. The classification between the groups of elderly people revealed that the combination of the easy dual-task and the fast walking task had the best prediction performance (AUC = 0.86). Gait variability in step and stride time and the RMS value of vertical acceleration were the features with the largest predictive power. SSC prediction accuracy was better than the accuracies obtained with linear discriminant analysis and support vector machine classifiers.

**Conclusions:** The study corroborated that the changes in gait patterns can be used to discriminate between young and older adults and more importantly between healthy and cognitively impaired adults. A subset of gait tasks and STGF optimal for achieving this goal with SSC were identified, with the latter method superior to other classification techniques.

## 1 Introduction

Human gait patterns naturally change across the lifetime, adapting to changes in structural and muscular characteristics. The decrease in muscle mass, and aging of the bones and joints, among other musculoskeletal changes, produce a limited range of motion, the avoidance of weight-bearing, asymmetry, and limping in elderly persons (Pirker and Katzenschlager, 2017). Thus elderly adults walk more slowly than the youths, with a shorter step length and increased time in double limb support (Menz, Lord and Fitzpatrick, 2003; Lønseth, 2016), among other changes. Gait disorders have a prevalence of 10% between 60 and 69 years, this increases to 60% in adults over 80 years (Mahlknecht *et al*., 2013). These impairments produce recurrent falls, depressed mood, and diminished quality of life (Mahlknecht *et al*., 2013; Montero-Odasso *et al*., 2012). The causes of gait disorders include neurological conditions, where cognitive impairments play a central role. Interestingly, gait disturbances in older adults with apparently normal cognition could predict early cognitive decline (Buracchio *et al*., 2010; Bahureksa *et al*., 2016; Byun *et al*., 2018; Beauchet *et al*., 2013; Kikkert *et al*., 2017).

The performance of spatio-temporal gait features (STGF) has been explored with different walking tasks in healthy and cognitively impaired adults. Dual-task, fast, and normal walking tasks, are the tasks most frequently reported in the literature. The dual-task paradigm has become the preferred test for assessing interactions between cognition and gait, in addition to the risk of falls (Lundin-Olsson, Nyberg and Gustafson, 1997). In this paradigm, the STGF obtained when the subject performs an attention-demanding task and walks at the same time are compared with normal walking (Pashler, 1994). The premises are that two simultaneously performed tasks compete for brain resources (Yogev-Seligmann, Hausdorff and Giladi, 2008; Montero-Odasso *et al*., 2009), and thus interference is greater when these resources are reduced (e.g. by brain damage). Some studies find that dual-task paradigms are good predictors of cognitive decline in healthy older adults (MacAulay *et al*., 2017; Beauchet *et al*., 2017), and also predict well progression to dementia in patients with Mild Cognitive Impairment (MCI) (Montero-Odasso *et al*., 2017) Moreover, they allow discrimination between cases of Parkinson’s disease (PD) that have and do not have MCI (Ricciardi *et al*., 2020) However, other studies find that walking at a fast pace is enough to increase sensitivity for MCI diagnosis (Bahureksa *et al*., 2016; Beauchet *et al*., 2013). Lønseth (2016) report that walking at a preferred pace is sufficient for predicting cognitive decline from STGF. In addition, Ricciardi et al. (2020) propose that normal gait could be used for screening MCI in PD due to its high sensitivity. As can be seen, there is no agreement regarding the most appropriate walking condition to assess the relationship between gait and cognitive impairment (Bahureksa *et al*., 2016).

On the other hand, several methodological advances facilitate the use of gait analysis in cognitive assessments. First, inertial measurement units (IMU) enable gait analysis outside laboratories and during long periods (Benson *et al*., 2018; Gwak, Sarrafzadeh and Woo, 2018; Chen *et al*., 2020; Mannini *et al*., 2015; Dasgupta, Vanswearingen and Sejdic, 2018). Second, machine learning techniques (MLT) allow extracting more information from the data and uncovering unforeseen relationships between variables (Graham *et al*., 2020). A brief review illustrates these possibilities. Gwak, Sarrafzadeh and Woo (2018) applied a logistic regression classifier to photoplethysmography and STGF and reported an 86% identification accuracy of MCI. Chen et al. (2020) explored several classifiers based on STGF for discriminating cases of Parkinson’s disease with MCI and without MCI. They found that feature selection, based on principal component analysis (PCA), followed by classification with a support vector machine (SVM) using a polynomial kernel was the most accurate classifier (91.67%). Another study (Zhou *et al*., 2020) employed SVM combined with PCA, random forests (RF), and Artificial Neural Network (ANN) on the STGF of normal walking in samples of young adults, healthy elderlies, and geriatric elderlies (with many comorbidities, but without cognitive impairment). They were able to classify the age accurately with AUC values of 0.91 (0.81– 0.93), 0.86 (0.63–0.83), and 0.86 (0.72–0.87) for each algorithm. Ghoraani et al. (2021) applied SVM classifiers based on STGF and Montreal Cognitive Assessment (MoCA) for the following discriminations: MCI vs. healthy, Alzheimer’s disease (AD) vs. healthy, and AD vs. MCI. They obtained average accuracies of 78% (77% F1-score) and 83% (84% F1-score) with STGF and MoCA scores, respectively. Dasgupta, Vanswearingen and Sejdic (2018) were able to accurately predict small changes in gait patterns due to cognitive load in a small sample of healthy adults using either logistic regression, RF, learning vector quantization, or SVM.

Unfortunately, most attempts to select the optimal experimental/STGF paradigms have compared only a few variants of walking tasks and STGF combinations, and only in small subject samples. This entails slow progress in defining the relative merits of the many potential combinations of tasks/features that exist. Consequently, there are still no standardized clinical tools for predicting the cognitive status from STGF, with a great variety of experiments and variables waiting to be explored. Solving this with traditional approaches would imply many experiments, each ideally with a large sample of subjects (with many more cases than the number of STGF). The Stable Sparse Classifiers procedure (SSC) proposed Bosch-Bayard et al. (2018) could help overcome this limitation because it can work with samples of a size greater than or similar to the number of STGF, allowing different combinations of these features to be obtained without having to increase the sample size.

SSC is applied to find more robust biomarkers by using resampling techniques. SSC estimates a penalized regression model and computes the precision and stability of the selected biomarkers. The procedure uses the glmnet package of MATLAB for estimating the penalized regression (Hastie and Qian, 2016). It then applies nested cross-validation to the data. Glmnet has been previously used for the development of a graphical representation of a mathematical function, called a nomogram, that predicts cognitive impairment in healthy older adults from different risk factors (including mobility) (Liu *et al*., 2021). SSC helps to deal with the fact of having similar numbers of features and subjects (Bosch-Bayard *et al*., 2018), and thus it is an ideal candidate to apply to our study.

Here we tested if SSC based on STGF could discriminate between young adults and healthy elderly adults (as a proof of concept) and more importantly if this approach could discriminate between healthy and minimally cognitively impaired older adults. We recorded gait patterns using IMU with a moderate-sized sample of participants. Participants were assessed with 5 tests from different cognitive domains. A summarized cognitive index (MDCog), based on the Mahalanobis distance from normative data, was calculated. In another study (Aznielle *et al*., 2022), we have demonstrated that MDCog has a closer relationship with STGF than other cognitive measures. An additional goal was to use SSC to identify the best combination of walking tasks and associated STGF to predict cognitive status.

## 2 Materials and Methods

### 2.1 Participants

Participants were selected from different places of Havana city: the Cuban Center for Neuroscience (CNEURO), three nursing homes, several grandparent homes, and primary health care areas. The inclusion criteria were the participant’s agreement, with ages between 20 and 40 years for young adults and above 60 years for older adults, and a Katz Index of independence ≥ 4 (Shelkey and Wallace, 2000). This index measures functional independence, based on the need for supervision or external help in performing basic activities of daily life (Katz *et al*., 1963). Participants were excluded if they could not walk or had to use walking aids, presented major neurological disorders, diseases of the musculoskeletal system, or severe cognitive impairment that prevented complying with instructions. Before gait evaluation, all participants underwent a neurological examination, filled out a questionnaire designed to explore the participant’s habits, and underwent a cognitive assessment explained in the following section. Ultimately, 125 participants completed all the requirements and participated in the study, divided into 40 young adults (mean age 27.65 ± 4.14, 50% women) and 85 older adults (mean age: 73.25 ± 6.99, 62.3% women). Written informed consent was obtained from the participants or caregivers and the study was approved by the ethics committee of CNEURO, which verified compliance with the Helsinki declaration.

### 2.2 Cognitive assessment

Five neuropsychological tests were applied for assessing the participant’s cognitive trait: Mini-Mental State Examination (MMSE), Attentional Span or Brief Test of Attention (BTA), Trail Making Test (TMT), Hopkins Verbal Learning Test (HLVT), and Digit Symbol Substitution Test (DS). Global cognition was assessed by using the MMSE (Folstein, Folstein and McHugh, 1975). BTA is a measure of auditory divided attention (Schretlen, Bobholz and Brandt, 1996). Attention, visuospatial abilities, mental flexibility, and executive functions were assessed by the TMT, parts A and B (Rabin, Barr and Burton, 2005). Memory was evaluated by HLVT, including immediate recognition and delayed recall (Brandt, 1991). The DS evaluated the focused, selective, and sustained attention as well as visual perception (Shum, McFarland and Bain, 1990). Twelve cognitive measures were extracted. Normative data for these tests had been obtained for the Cuban adult population in an international collaborative study (Rivera *et al*., 2015; Arango-Lasprilla, Rivera, Aguayo, *et al*., 2015; Arango-Lasprilla, Rivera, Garza, *et al*., 2015; Arango-Lasprilla, Rivera, Rodríguez, *et al*., 2015).

### 2.3 Group definition

The cognitive index MDCog was calculated for each participant (Aznielle *et al*., 2022). This cognitive index is an objective and continuous measure of the subject’s cognitive status. It takes into account the results in each neuropsychological test, adjusted by age and educational level, and compensated for the correlation between them. It summarizes the overall deviation of the subject’s cognitive performance from the normative data. The participants were classified into two groups (healthy and impaired), using k-means clustering of MDCog (1000 repeats). The healthy group was split based on age. Thus three groups were defined: young adults (YA), healthy older adults (HE), and cognitively impaired older adults (MCI-E).

### 2.4 Walking tasks

Participants executed four walking tasks in an obstacle-free and flat environment. They covered 40 m (20 m in one direction and 20 m in the opposite direction). The walking tasks were: 1) walking freely at a comfortable self-chosen speed (NormalW); 2) walking at a comfortable self-chosen speed while simultaneously counting their steps, an easy cognitive task (EasyD); 3) walking at a comfortable self-chosen speed while simultaneously counting backward from 100, a hard cognitive task (HardD); and 4) walking as fast as possible without running (FastW). Counting backward is hard because it requires both working memory and attention (Montero-Odasso *et al*., 2009). Participants were not instructed to prioritize walking or counting.

### 2.5 Gait assessment

Data were collected with the Bitalino RIoT (Plux Wireless Biosignals, Portugal) IMU, attached to a velcro band, and worn firmly near the body’s center of mass, on the lower back at L3 spinal level. The software OpenSignals (Plux Wireless Biosignals, Portugal) was used to record the data. The IMU data axes were transformed to comply with the International Society of Biomechanics (ISB) recommendations (Wu and Cavanagh, 1995).

The recorded gait patterns were split into two segments, one for each direction of the walk. The strides corresponding to the first and last three seconds of each segment were removed. Dynamic tilt correction (Moe-Nilssen, 1998; Millecamps *et al*., 2015) was applied to compensate for the imprecise position of the IMU and the effects of gravity on the measured accelerations. Acceleration signals were low-pass filtered (4th order zero-lag Butterworth filter at 3 Hz). The vertical acceleration signal was used since it is more robust and greater reliable than the two other axes (Alvarez *et al*., 2006; Maquet *et al*., 2010; Kavanagh *et al*., 2006).

The initial contacts (IC) and final contacts (FC) instants of the gait cycle were estimated with the method proposed by McCamley et al. (2012). This method smooths the vertical acceleration signal by integration (Zok, Mazzà and Della Croce, 2004), and then obtain two derivatives using a Gaussian continuous wavelet transform (Luo, Bai and Shao, 2006). The ICs corresponded to the minima of the first derivative. The FCs correspond to peaks in the second derivative. Then sixteen STGF were estimated: 1) step time (StpT); 2) step time variability or step time coefficient of variation (StpTCoV); 3) stride time (StrT); 4) stride time variability or stride time coefficient of variation (StrTCoV); 5) cadence (Cd); 6) root mean square amplitude of the vertical acceleration (RMS); 7) double support duration or double support time (DSD) (Jarchi <i>et al.</i>, 2018); 8) single support duration or single support time (SSD); 9) swing duration feed 1 (SwDurF1); 10) swing duration feed 2 (SwDurF2); 11) stance duration feed 1 (StDurF1); 12) stance duration feed 2 (StDurF2); 13) step duration feed 1 (StepDurF1); 14) step duration feed 2 (StepDurF2); 15) step length (StepLg); and 16) speed (GS). Almost all STGF are expressed in seconds (s), except Cd (steps/min), RMS (g), StepLg (m) and GS (m/s). STGF were computed using algorithms described in literature (Zijlstra, 2004; Del Din, Godfrey and Rochester, 2016; Montero-Odasso *et al*., 2011; Yang *et al*., 2012; Jarchi *et al*., 2018; Bugané *et al*., 2012; Zijlstra and Hof, 2003; Del Din *et al*., 2016).

Thus 64 STGF were computed. Dual-task costs (DTC) for the STGF in the two dual-tasks were also calculated (32 costs). These costs (expressed as percentages) were calculated using as follows (Montero-Odasso *et al*., 2017):

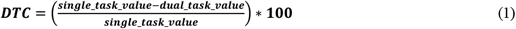

Finally, 96 measures (STGF and DTC) were obtained for each walking direction.

### 2.6 Stable Sparse Classifiers procedure

SSC consists of the following steps (Bosch-Bayard *et al*., 2018):

1. Biomarkers selection: Initially data is perturbed (70% of subjects and 70% of variables are randomly selected by resampling method), followed by a screening step for eliminating variables with a minimal contribution. Later, a smaller set of biomarkers is selected using glmnet (see Model formulation below) with cross-validation. These steps are repeated iteratively (n = 500) to identify the variables that have a frequency of selection above a consistency threshold (50%). The parameters of the models were calculated intrinsically by the procedure.
2. Model validation (using the variables selected previously) based on ROC and stability assessments: A random subsample with the 70% of the subjects is used to classify using glmnet, and the rest of the sample is used to calculate the ROC values. This procedure is executed in several iterations (n = 500) to estimate the distribution of ROC values. The AUC at 50 percentile of the distribution is used to measure classification accuracy, and the model with highest accuracy is then selected as the best model.

Model formulation: glmnet

The elastic-net model, used to select the biomarkers, is formulated as a weighted multivariate linear regression model, described by the equation (Zou and Hastie, 2005; Friedman, Hastie and Tibshirani, 2010):

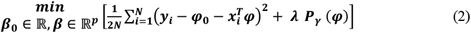

Where: N is the number of subjects, *x*_*i*_ ∈ ℝ^*p*^ observations of subject i, and *y*_*i*_ ∈ ℝ is the label group of subject *i*; *φ*_0_ ∈ ℝ, *φ* ∈ ℝ^*p*^are the model parameters; *γ* is the regularization parameter; *p* is the number of variables in the model; and:

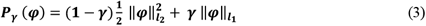

is the penalty equation known as elastic-net norm (Zou and Hastie, 2005). The use of L2 norm 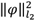 induces a regression (known as ridge regression) that behaves well for high dimensional data but tends to spread out coefficient weights among highly correlated variables, and L1 norm 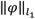 produces the “lasso regression”, indifferent to highly-correlated predictors, which tries to estimate only a few nonzero coefficients (i.e. performs variable selection), thus inducing sparsity in the vector of coefficients. *γ* and *λ* are the parameters of the relative contributions of the ridge and the lasso to the elastic net.

The characteristics of the elastic net allow SSC to deal with highly correlated features. In addition, the glmnet classification algorithm can solve the problems related to the high number of features and the low number of subjects. Both advantages are useful for this study.

### 2.7 Analysis

The analysis flowchart of this work is shown in **Figure 1**. The goals of this work are presented in section A of the **Figure 1**, and the corresponding analyses to achieve them are shown in section B. Since the 96 STGF from the two walking directions were highly correlated only their average was used.

**Figure 1.**
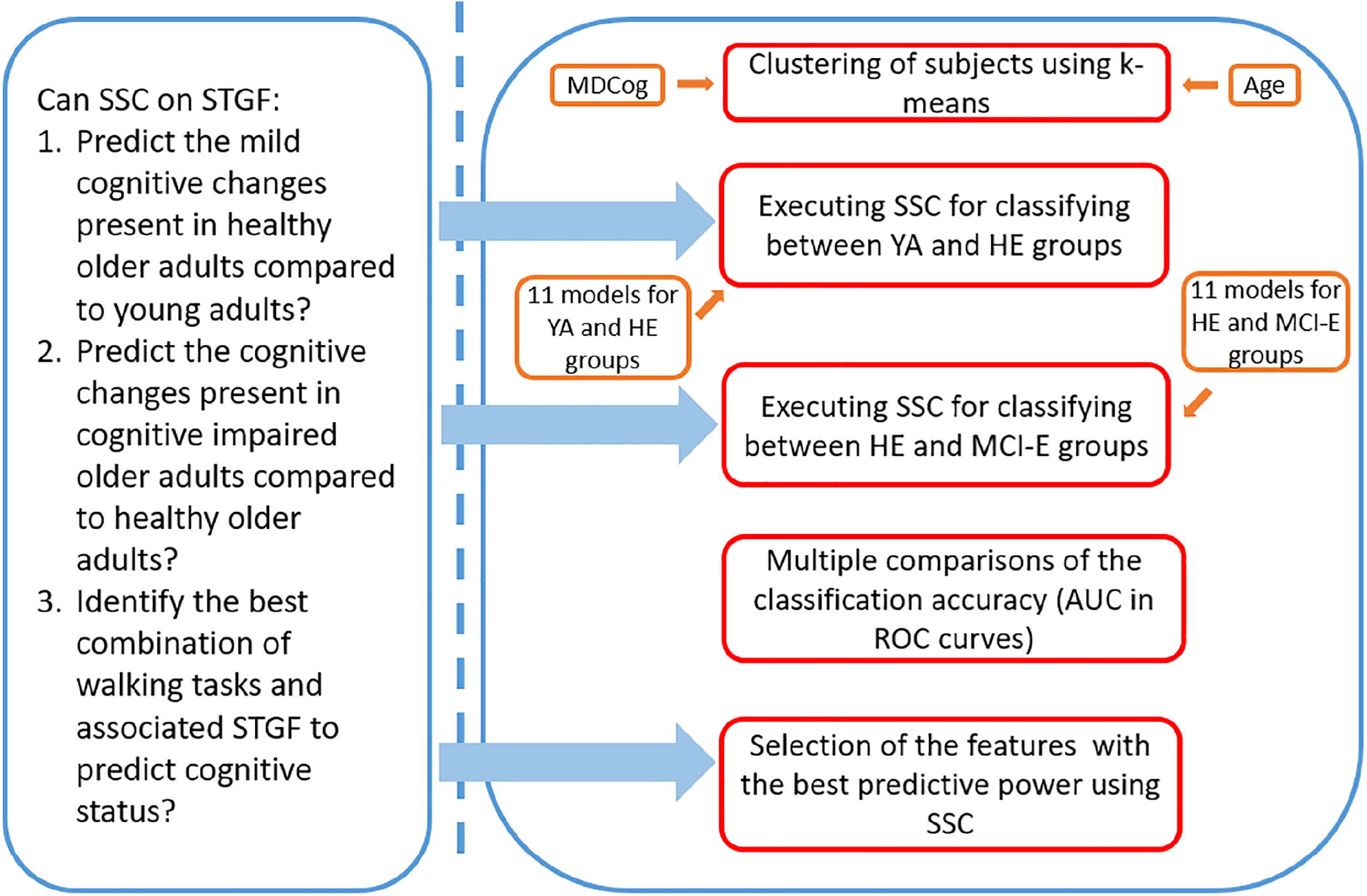
Analysis flow aimed to address the objectives of the work. **(A)** Questions derived from the goals of the study. **(B)** General description of the methods used to answer the questions and their interrelations.

SSC was applied for discriminating between: a) YA and HE (used as a sanity check), and b) HE and MCI-E (the main purpose of this study). In both cases, the following subsets of features (models) were explored: i) the STGF from all walking tasks lumped together (All-tasks); ii) the STFG for each walking task separately (e.g. NormalW); iii) the six combinations of STFG from pairs of walking tasks (e.g. NormalW+EasyD).

After applying SSC for the discrimination between HE vs. MCI-E, the Bonferroni pairwise comparisons test were performed to carry out post-hoc comparisons between the AUC of all models. SSC was applied also to identify the most stable variables in the two models with the highest accuracies.

The prediction accuracy of SSC was compared with two other classifiers: SVM and a regularized linear discriminant analysis (LDA).

## 3 Results

### 3.1 Clustering of subjects

The cognitive index MDCog ranged from 1.57 (the best performance) to 12.15 (the worst performance). The three clusters or groups obtained are presented in **Figure 2** and their demographic characterization is presented in **Table 1**. Note in the figure a slight overlap between the values of MDCog between the HE and MCI-E, with the cut-off at about 6. No differences in weight and height (which are known to affect gait) were found between the two elderly groups in a Kruskal-Wallis test. The elderly groups differed significantly from the YA group in these variables.

**Table 1.** Demographic characterization of the groups obtained by k-means clustering based on MDCog.

Stable Sparse Classifiers predict cognitive impairment from gait patterns
|  | Young-Adult (40) | Healthy-Elderly (62) | MCI-Elderly (23) |
| --- | --- | --- | --- |
| **Age (years)** | 27.65 ± 4.14 [22-38] | 72.23 ± 6.59 [60-88] | 76.00 ± 7.43 [61-87] |
| **Sex (% females)** | 20 (50) | 37 (59.7) | 16 (69.5) |
| **MDCog** | 2.91 ± 0.97 [1.57-5.13] | 3.81 ± 1.08 [2.00-6.16] | 9.30 ± 1.65 [6.50-12.15] |
| **Height (cm)** | 167.97 ± 8.19 [154-185] | 161.57 ± 9.92 [142-183] | 159.69 ± 8.95 [146-175] |
| **Weight (kg)** | 70.20 ± 14.69 [41-98] | 68.47 ± 16.59 [37-117] | 64.13 ± 14.63 [38-92] |

Values are presented as mean ± STD. The range is given in square brackets

**Figure 2.**
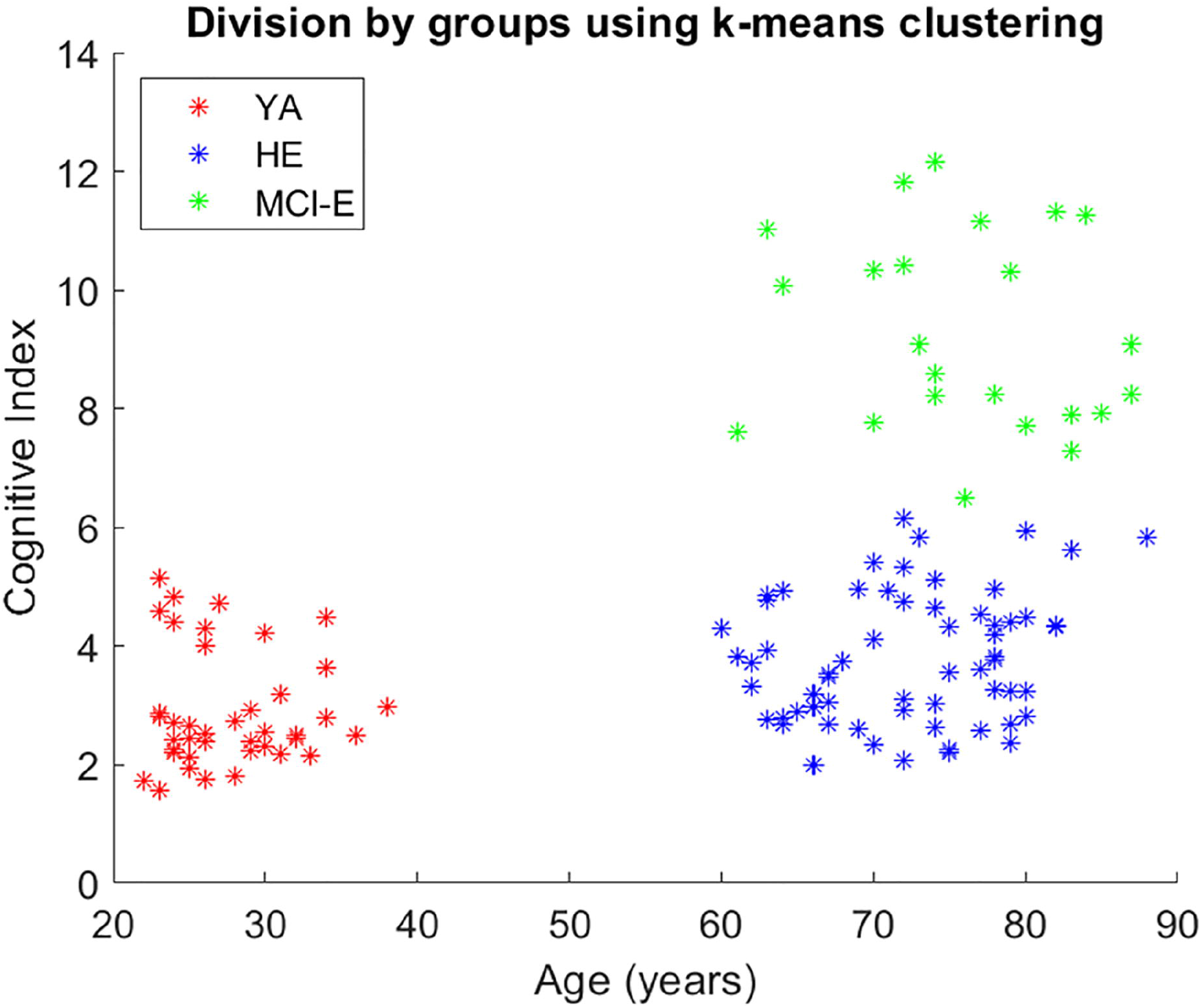
Division by groups using k-means clustering and Age variable.

### 3.2 Differences in STGF between groups

**Supplementary Figures 1** to **4** show the normalized values in regarding to mean and standard deviation (Z-scores) for each walking task in the three groups. The median and

The features related to the gait cycle phases and periods (i.e. gait variability and gait symmetry) were larger in the MCI-E group, somewhat smaller in the HE group, and even smaller in the YA. On the contrary, Cd, RMS, StepLg, and GS had larger values in the YA than in HE and MCI-E.

A non-parametric permutation two-way analysis of variance (ANOVA) test, with GROUP and TASK as main effects, was performed individually on each STGF. The interaction between these main effects was also tested. The results from this ANOVA are shown in **Supplementary Table 1**. For all STGF and all analyses, there were significant effects of GROUP and TASK (p < 0.001 in almost all cases. The only significant interaction effect found was for StepTimeVar (p < 0.02).

### 3.3 Discrimination between YA and HE

**Figure 3** shows the results of classification procedure for all defined models. The ROC curves on All-tasks and the individual walking tasks models are shown in **Figure 3A**. The same curves for walking task pairs are shown in **Figure 3B**. The AUC values ranged between 0.77 and 0.64. The models with the largest AUC were EasyD+FastW and All-tasks (both with 0.77), while the model with the smallest one was NormalW.

**Figure 3.**
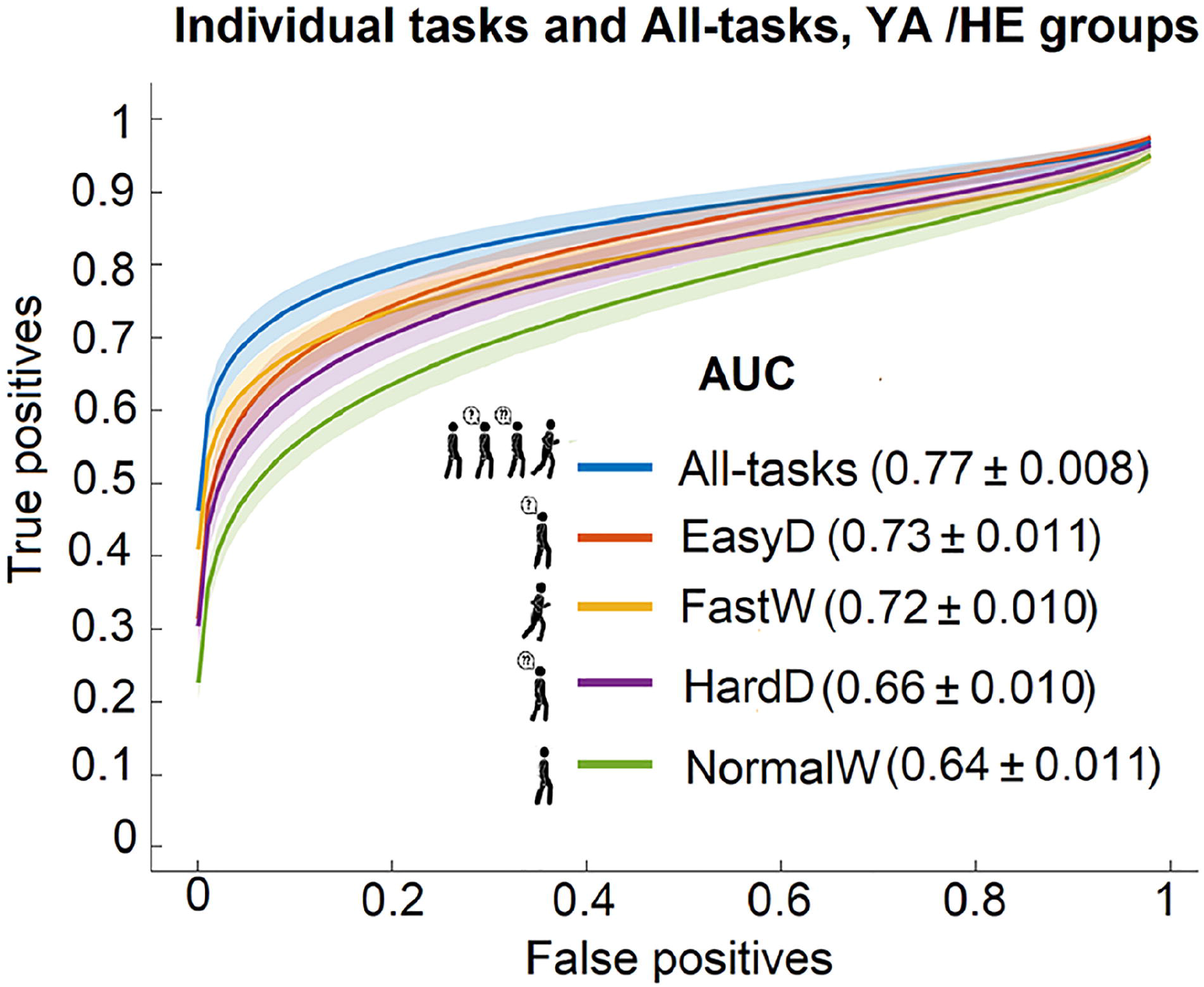
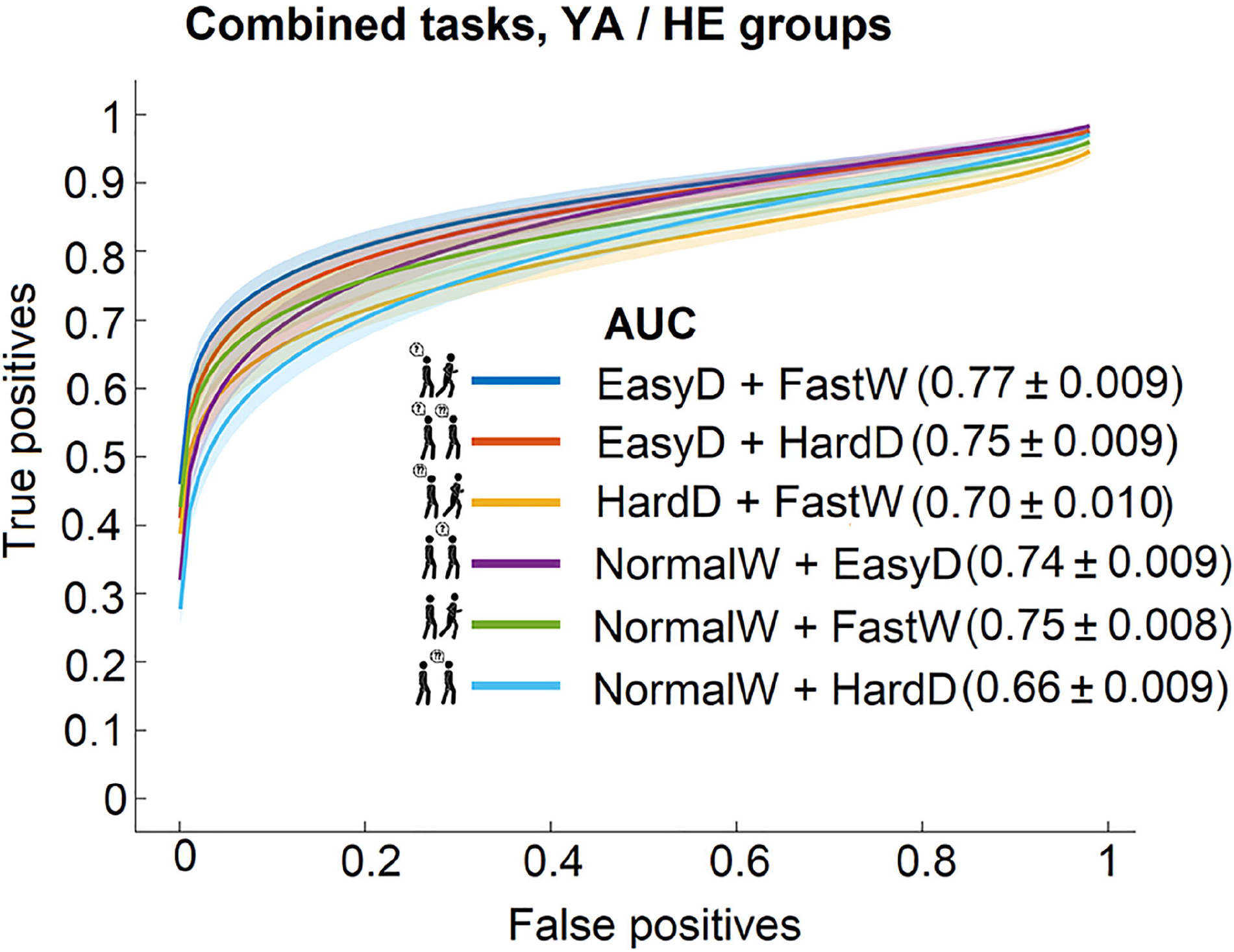
ROC curves with 95% confidence bands and model AUC values for predicting the cognitive status between YA and HE groups using **(A)** the individual tasks and the complete set, and **(B)** the combined tasks sets.

### 3.4 Discrimination between HE and MCI-E

The ROC curves for SSC between HE and MCI-E based on All-tasks and the individual walking tasks are shown in **Figure 4A**. The largest AUC was obtained with All-tasks, followed by FastW and EasyD tasks. Interestingly, the lowest AUC was obtained for NormalW and HardD tasks. **Figure 4B** shows the ROC curves for SSC between HE and MCI-E based on pairs of tasks. The task-pairs including FastW yielded the largest AUCs. The most accurate combination was SSC based on the EasyD+FastW combination (AUC = 0.86), which was close to that obtained with all the features together (All-tasks).

**Figure 4.**
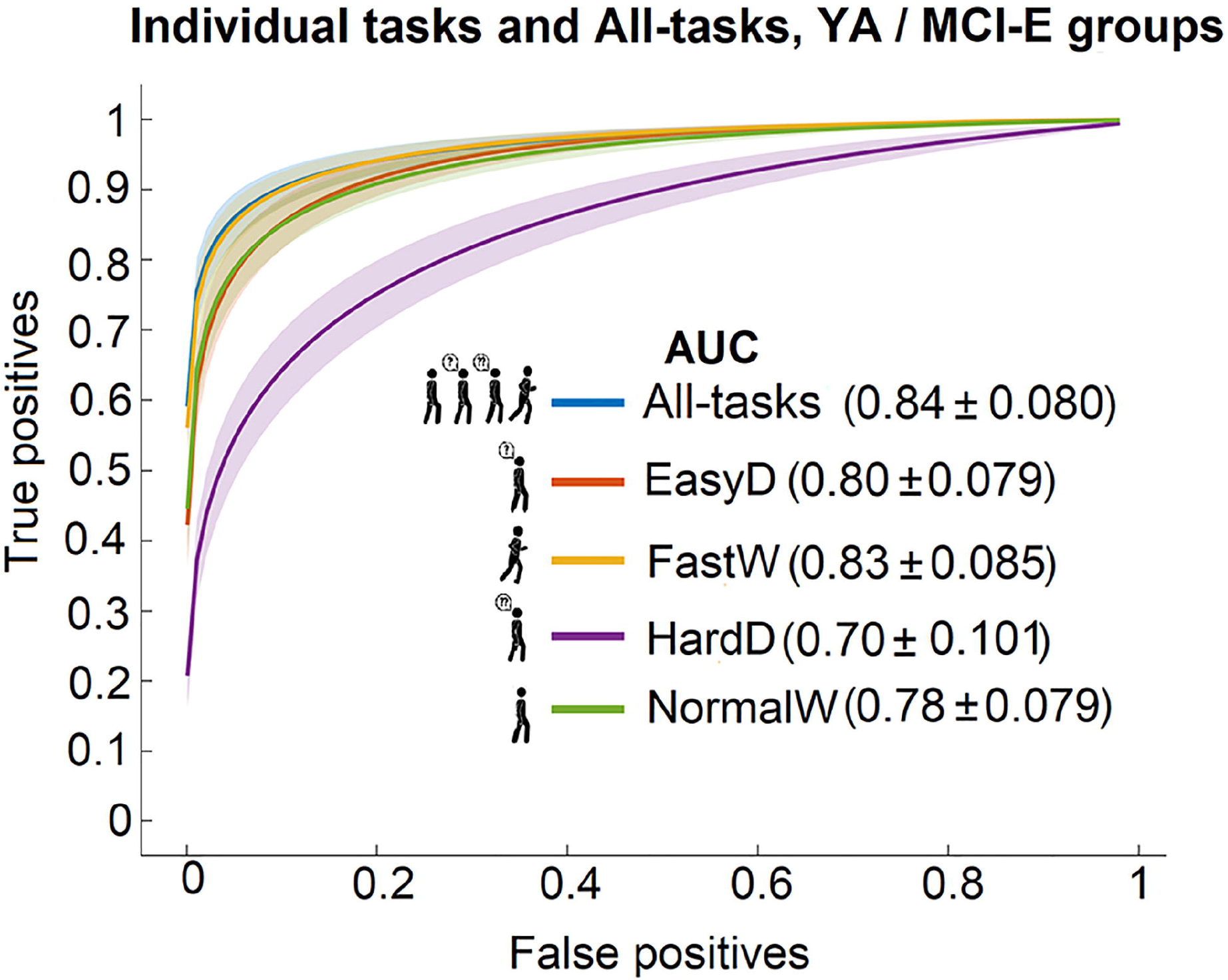
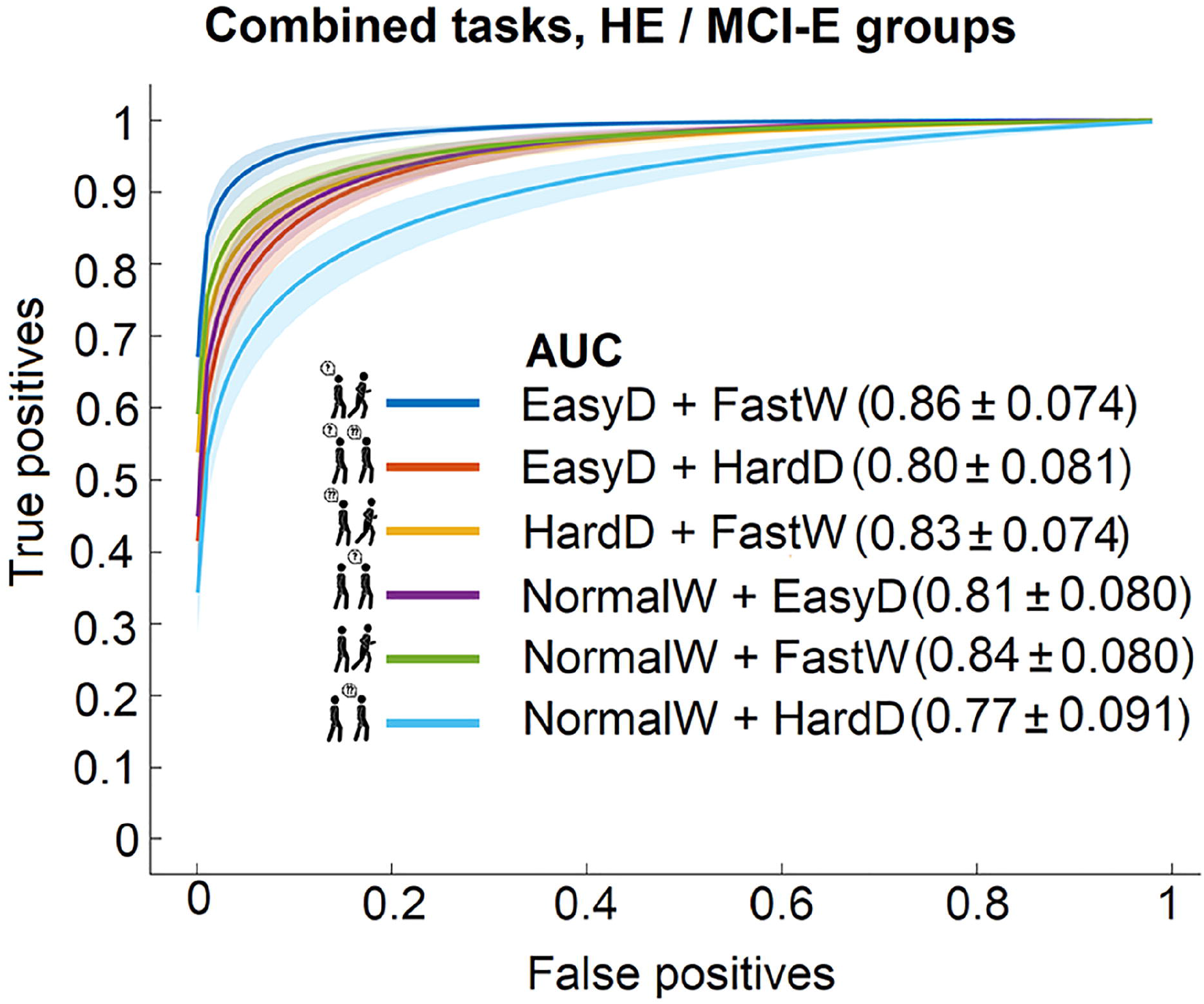
ROC curves with 95% confidence bands and model AUC values for predicting the cognitive status between HE and MCI-E groups using (A) the individual tasks and the complete set, and **(B)** the combined tasks sets.

**Figure 5** shows the results of a multiple-comparison procedure using NormalW model as a reference. The mean AUC value and the confidence interval for each model are presented. The AUCs for NormalW+HardD and EasyD+HardD models were not significantly different from NormalW (est = 89.58, p < 1.000 and est = -107.79, p < 0.904, respectively). The rest of the combinations were significantly different. The models with larger performance were NormalW+FastW and EasyD+FastW (est = -658.25, p < 0.0001 and est = -771.32, p < 0.0001, respectively). The only model that underperformed with respect to the reference was HardD (est = 208.00, p < 0.0002).

**Figure 5.**
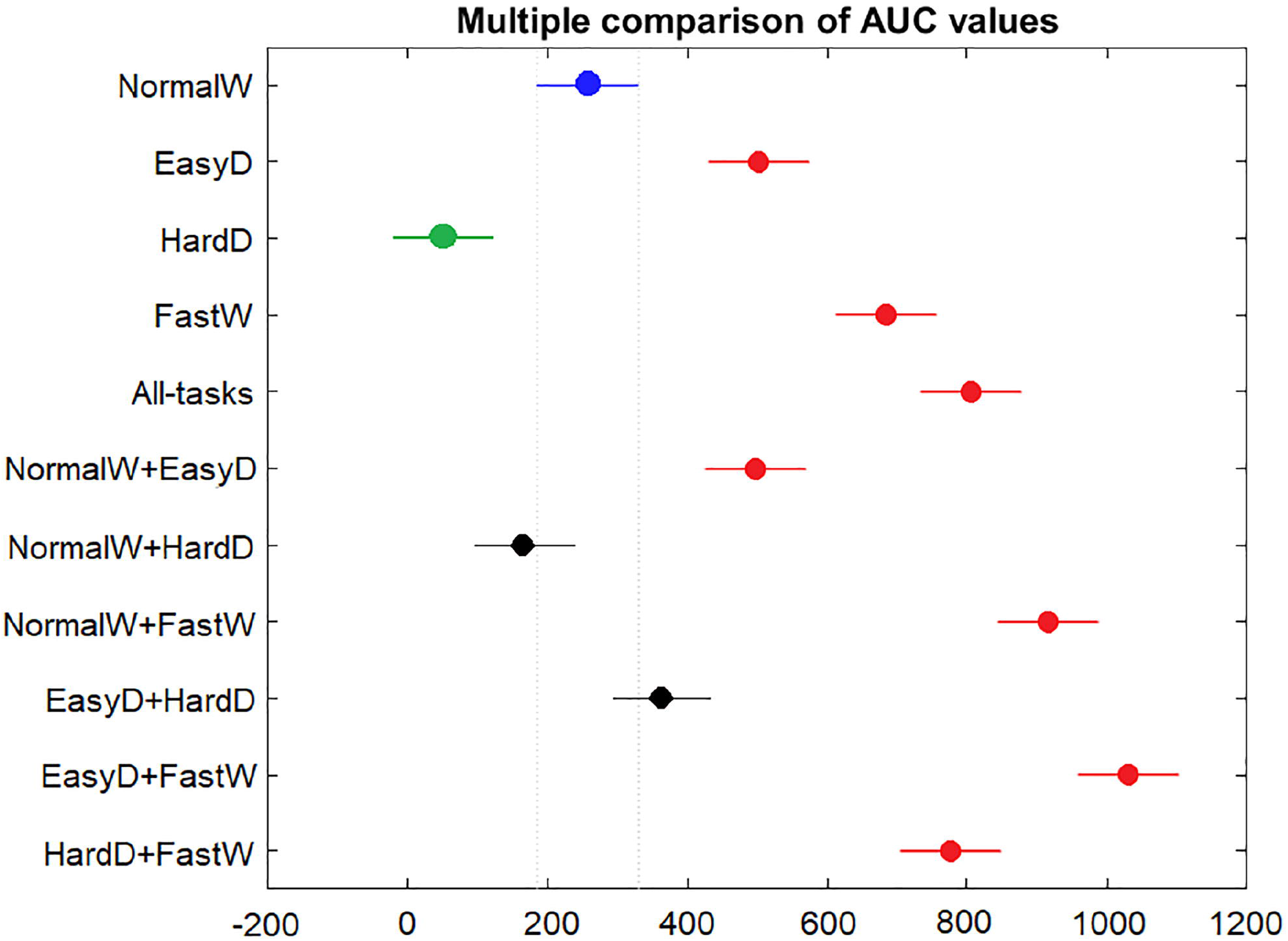
Estimates and comparison intervals (Bonferroni) of the AUC values in individual and combined by pair tasks between the two groups of elderlies, calculated by the multcompare function of Matlab. The blue marker is the reference task, black dot indicates no difference with this reference, red dots mean more significance from the reference, and green ones, less significant than the reference.

### 3.5 Contribution of selected stable biomarkers to the two best models

The selected biomarkers in the best models for discriminating between HE and MCI-E (NormalW+FastW and EasyD+FastW tasks) were analyzed. **Figure 6** shows a plot with the proportions of time that each STGF was retained in SSC.

**Figure 6.**
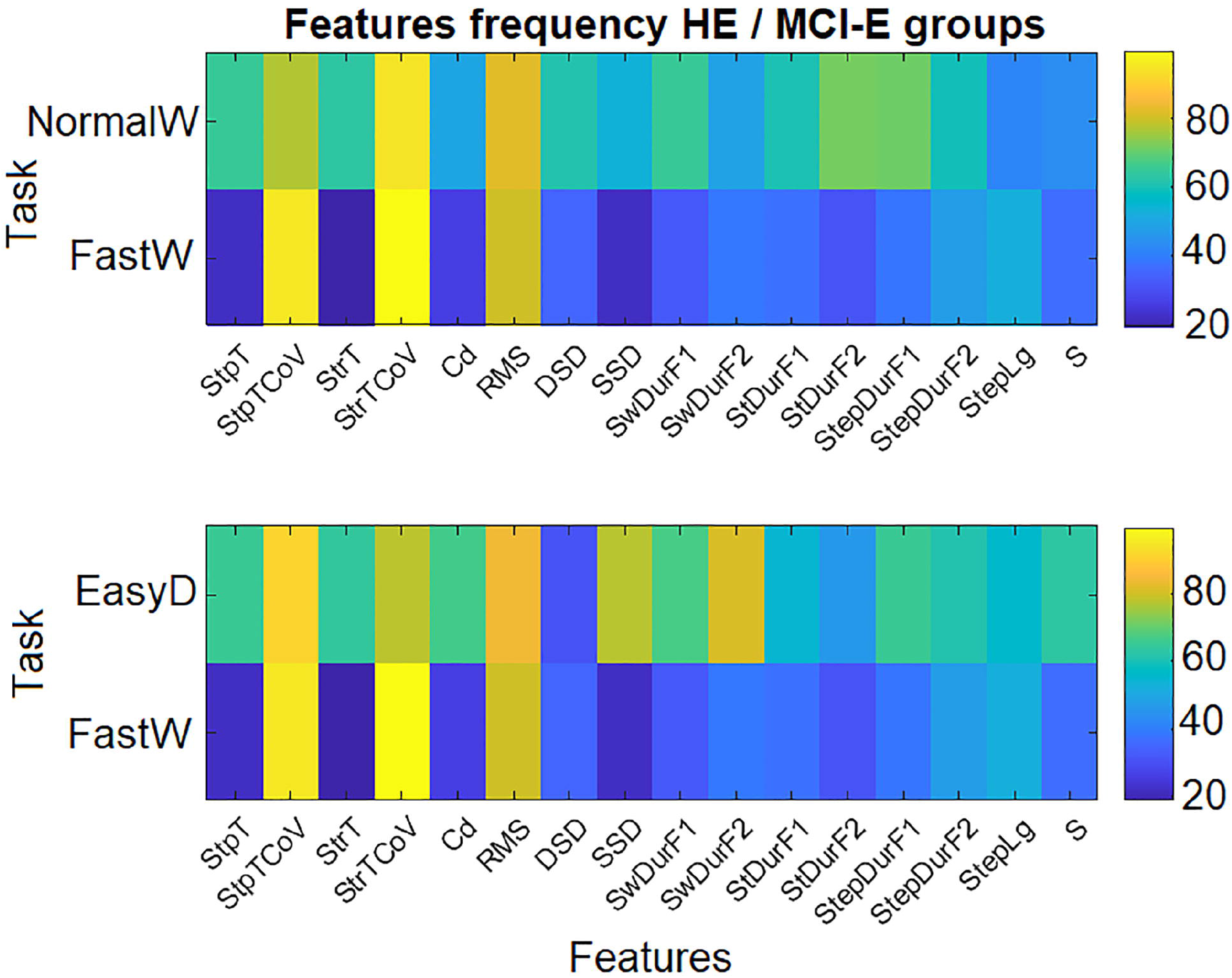
The proportion of time (%) each measure appeared as significantly for two estimated models across iterations: **(A)** NormalW+FastW and **(B)** EasyD+FastW.

The best predictors in both models belonged to the FastW task. Specifically, they were the gait variability in step and stride times (StpTCoV and StrTCoV) and the RMS value. Also, the same features in NormalW and EasyD tasks were selected in each model, respectively.

Other features related to the description of the gait cycle (NormalW: StDurF2 and StepDurF1; EasyD: SSD and SwDurF2; HardD: SwDurF1) were also selected with high predictor power. In correspondence to wahta was explained in section 3.2, the features related to gait variability and gait cycle had higher means in the MCI-E group than in HE group. Contrary, the RMS values were higher in the HE group as expected result (see **Supplementary Figures 1** to **4**).

### 3.6 Comparison of SSC performance with other classifiers for discriminating HE and MCI-E

The means and standard deviations of AUC values for classifications between HE and MCI-E with SSC, LDA and SVM are presented in Table 2. As shown in this table, the best results were achieved with SSC. The other two classifiers had lower performances in all models. There were significant differences between the AUC of the three classifiers. Interestingly, the EasyD+FastW SCC with AUC of 0.86 was more accurate than SVM (AUC = 0.76) and LDA (AUC = 0.83).

**Table 2.** AUC values for the three classifiers using the eleven models for HE and MCI-E groups.

| Model | SSC | SVM | LDA |
| --- | --- | --- | --- |
| **NormalW** | 0.78 ± 0.08* | 0.67 ± 0.04* | 0.62 ± 0.06* |
| **EasyD** | 0.80 ± 0.09* | 0.75 ± 0.01* | 0.69 ± 0.06* |
| **HardD** | 0.70 ± 0.1* | 0.52 ± 0.04* | 0.59 ± 0.03* |
| **FastW** | 0.83 ± 0.09* | 0.72 ± 0.03* | 0.79 ± 0.03* |
| **All-STGF** | 0.84 ± 0.08* | 0.59 ± 0.04 | 0.65 ± 0.05 |
| **NormalW+EasyD** | 0.81 ± 0.08* | 0.74 ± 0.02* | 0.65 ± 0.07* |
| **NormalW+HardD** | 0.77 ± 0.09* | 0.64 ± 0.04* | 0.53 ± 0.03* |
| **NormalW+FastW** | 0.84 ± 0.08* | 0.69 ± 0.03* | 0.73 ± 0.04* |
| **EasyD+HardD** | 0.80 ± 0.08* | 0.70 ± 0.08* | 0.60 ± 0.02* |
| **EasyD+FastW** | 0.86 ± 0.07* | 0.76 ± 0.03* | 0.83 ± 0.03* |
| **HardD+FastW** | 0.83 ± 0.07 | 0.62 ± 0.03* | 0.82 ± 0.04 |

\* significant differences

## 4 Discussion

The relationship between STGF (measured in four walking tasks with IMU sensors) and the cognitive status of elderly and young participants was studied here. All the STGF differed significantly between groups and walking tasks, with no interactions between these effects. Accurate classification of the participants into our experimental groups was found with SSC. The best SSCs discriminating between healthy and cognitively-impaired adults were obtained with a combination of EasyD and FastW tasks. Contrary to our expectations, classification based on the HardD task was poor. Variability of step and stride time, and the RMS value of vertical acceleration were the features that contributed most to classification power. Compared to SVM and LDA, the SSC was more accurate in classifying the participants into their groups.

We do not need sophisticated gait measurements to distinguish YA from HE. However this comparison serves as a proof of concept for our methods. Similar to many other studies we found statistically significant differences in STGF between YA and HE results (Menz, Lord and Fitzpatrick, 2003; Zijlstra, 2004; Kobsar *et al*., 2014). Part of these effects related to aging could be due to impairments in skeletal-muscular function, but small alterations in brain function could also be playing a role. Previous work has been able to adequately discriminate YA and HE with other classifier algorithms. Previously, Begg and Kamruzzaman (2005) were able to discriminate between YA and HE, based on STGF extracted from normal walk, using SVM with different kernel functions. The got accuracies between 0.58 and 0.62 for the different kernels, similar values to our accuracy with the STGF from NormalW (0.61).

More relevant was the comparison between HE and MCI-E since the latter condition may not be so obvious to detect. We found statistically significant differences in all STGF between the HE and MCI-E groups. Previous articles (using smaller sets of STGF than ours) describe similar results to this when comparing older adults that are healthy and those with different degrees of cognitive impairment (Buracchio *et al*., 2010; Kikkert *et al*., 2017; MacAulay *et al*., 2017; Beauchet *et al*., 2017; Montero-Odasso *et al*., 2017; Zijlstra, 2004) or with neurological pathologies (Ren *et al*., 2017; Fitzpatrick *et al*., 2007). The fact we did not find interactions between GROUP and TASK in our ANOVAs indicates that alterations in gait patterns were pervasive and showed up in all tasks. Thus, we replicate previous findings while extending them to a much larger number of STGF.

In partial agreement with previous studies, we found that STGF from dual walking tasks improved discrimination of SSC. The FastW and EasyD tasks, alone and in combination, predicted cognitive status very well (although the HardD did not). To our knowledge, the use of counting steps aloud has not been previously reported as a walking dual-task. Fast walking has been previously described as more demanding than normal walking, and consistent with this, subjects with cognitive impairment (even in its early stages) perform it worse (Bahureksa *et al*., 2016; Montero-Odasso *et al*., 2011; Fitzpatrick *et al*., 2007). Reduced speed for fast walking is a sensitive predictor of impaired cognitive function (Bahureksa *et al*., 2016; Beauchet *et al*., 2013; Callisaya *et al*., 2017). Here we found that combining both EasyD+FastW enhanced prediction of cognitive impairments.

Gait quality should deteriorate with increased dual-tasks difficulty, especially when there is cognitive impairment (MacAulay *et al*., 2017; Montero-Odasso *et al*., 2009). However, here STGF from the HardD task (count backward from 100) did not enable better discrimination between HE and MCI-E compared to STFG from the EasyD task (count steps). The opposite result was found. One explanation for this is that perhaps not all of our MCI-E subjects understood the HardD task well, or were unable to perform it consistently. This could have increased the variability in STGF within the same subject, or between subjects, in the MCI-E group. For example, gait could momentarily improve if subjects abandon the HardD task for short periods of time, something we informally observed in many cases. Thus making the dual-task too hard may impair its diagnostic utility.

Interestingly, other authors have also reported low correlations between dual-task STGF (or dual-task costs) and cognitive performance, both in older adults (Lønseth, 2016; Kikkert *et al*., 2017) and in subjects with Parkinson’s disease (Gaßner *et al*., 2017). The diversity in sample selection, the lack of standard protocols for the dual-tasks (Montero-Odasso *et al*., 2012), and the peculiarities of task structure (Halvorson, 2013) have advanced as explanations for these negative results. The specification of the dual-task paradigm seems to be the most important issue for detecting cognitive impairment with gait measurements (Montero-Odasso *et al*., 2017). Thus failure in task compliance due to an excessively high cognitive load is the most probable explanation for the low classification accuracy we found using HardD STGF. More naturalistic tasks that would be easier to perform, such as listening to conversations of increasing complexity, should be explored to solve this problem.

Higher accuracies were found here for SSC compared to SVM and LDA classifiers. However, this may not only be due to the advantage of the glmnet over these other algorithms in handling many features with few subjects. In SSC, identification of stable sparse biomarkers is executed prior to building the classifier in the prediction stage. In addition, the methodology uses different cross-validation algorithms which ensures prediction stability and techniques to avoid overfitting (Bosch-Bayard *et al*., 2018). These additional steps should also be combined with other algorithms (e.g. SVM) for a fairer comparison. Also, only the default kernel was used with SVM. Other kernels could improve its performance.

Similar to previous studies, we found that features related to gait variability (StpTCoV and StrdTCov), and RMS, had the best predictive power. Previous work has reported that gait variability is larger in healthy older adults than young adults at a normal pace (Kobsar *et al*., 2014). Increased gait variability in the fast walk of older adults with mild cognitive impairment and Alzheimer’s disease, compared to healthy older adults, has also been reported (Beauchet *et al*., 2013). The reduction of walking speed in older adults has been associated with the reduced magnitude of their RMS (Menz, Lord and Fitzpatrick, 2003; Zhong, Rau and Yan, 2018; Kikkert *et al*., 2017).

This study has several limitations. One is that the sample of older adults with cognitive impairment is smaller than the sample of healthy older adults, which could affect the reliability of our findings. Moreover, the sample of healthy and cognitively impaired older adults was not completely paired by sex and age. This disbalance in the sample could affect any classifier used, not only SSC. Further work should examine the possibility of predicting cognitive scores (not only classifying into groups) through regression analysis, a goal for which preliminary results are presented in Aznielle et al. (2022). Also, SSC should be compared with other classifiers, not only SVM and LDA.

## 5 Conclusions

This study confirms that the changes in gait patterns can be used to discriminate between healthy and cognitively impaired adults. The use of SSC improves classification accuracy over traditional approaches with classifiers like SVM and LDA. The SSCs using STGF from combinations of EasyD and FastW tasks produced the most accurate results, and were equivalent to using all the features. Hence, it is possible to use a reduced battery of walking tasks resulting in lower testing times (which is important with elderly patients). Since this study was carried out with IMU in naturalistic environments, our results suggest that applications of the methods in clinical settings should be explored.

## Data Availability

All data produced in the present study are available upon reasonable request to the authors

## 6 Article type

This article is proposed as Type A.

### 6.1.1 Permission to reuse and Copyright

Copyright © 2022 Aznielle-Rodríguez, Ontivero-Ortega, Galán-García, Sahli and Valdés-Sosa.This is an open-access article distributed under the terms of the Creative Commons Attribution License (CC BY). The use, distribution or reproduction in other forums is permitted, provided the original author(s) or licensor are credited and that the original publication in this journal is cited, in accordance with accepted academic practice. No use, distribution or reproduction is permitted which does not comply with these terms.

## 8 Author Contributions

TA: gait recordings, data curation, software, and writing. MO: methodology, formal analysis, reviewing, and editing. LG: methodology, formal analysis, and reviewing. HS: conceptualization, methodology, formal analysis, reviewing, and editing. MV: conceptualization, methodology, formal analysis, reviewing, and editing.

## 10 Acknowledgments

We thank the participants in this study, as well as the older adult’s caregivers who gave their consent. To Daysi García Agustín, Ana Fernández Nin and Yoelvis Pozo Burgos for the design and execution of the neurological examination. To Karen Aguilar Mateu for the design of the neuropsychological test battery, and to Ana Castro Laguardia, Neisbet Blasco Fanego, Brenda Peón and José Aníbal Ojeda Núñez for the application and evaluation of the tests. To Jaime Menéndez Álvarez, Leisy Serrano Blanco and Gianna Arencibia for recording the gait patterns of the study participants. Lastly, we also acknowledge the technical assistance of Agustín Lage, Eduardo Martínez and Eduardo Garea.

