## Supplemental Material for "Stable Sparse Classifiers predict cognitive impairment from gait patterns"

Supplementary Material

### 1 Supplementary Figures and Tables

#### 1.1 Supplementary Figures

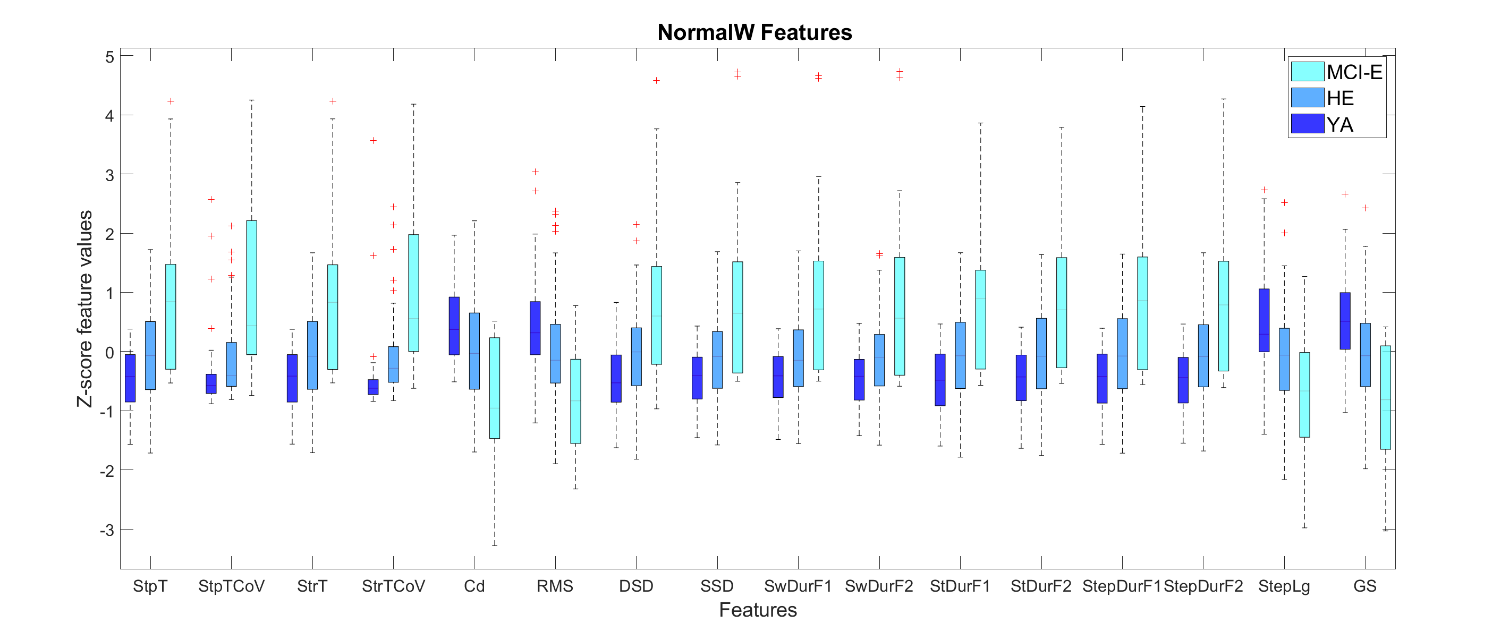

**Supplementary Figure 1.** Z-scores of STGF for the three groups in NormalW walking task.

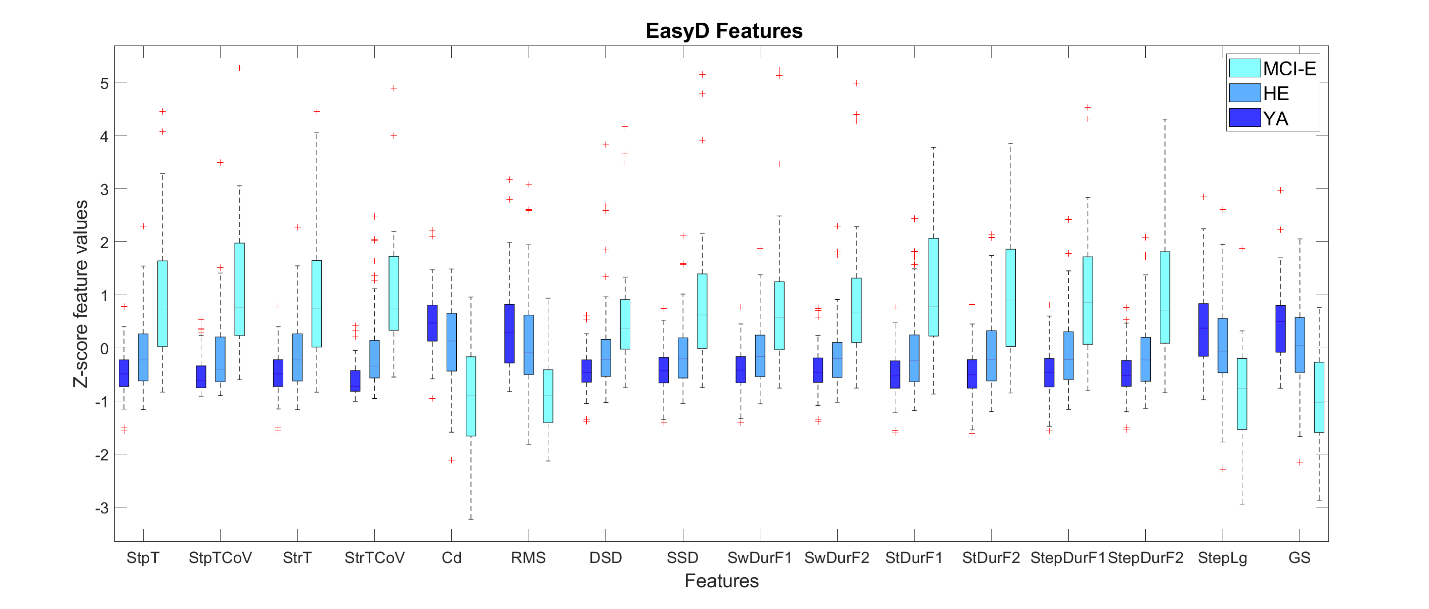

**Supplementary Figure 2.** Z-scores of STGF for the three groups in EasyD walking task.

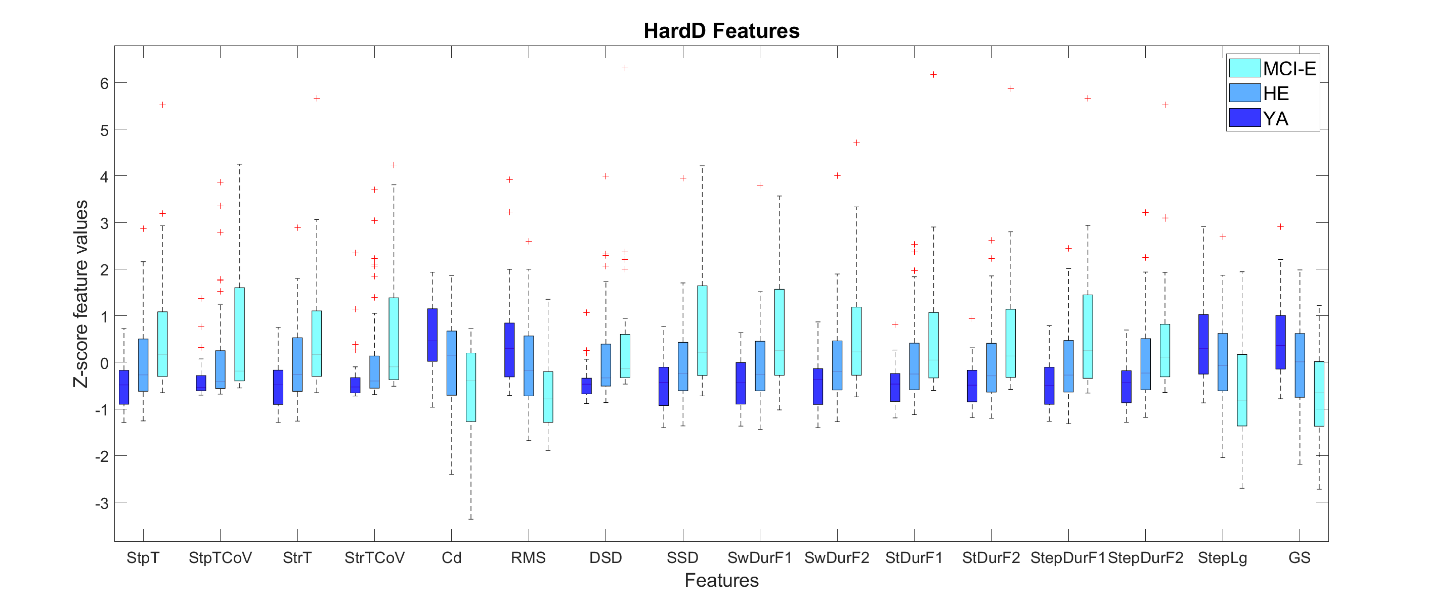

**Supplementary Figure 3.** Z-scores of STGF for the three groups in HardD walking task.

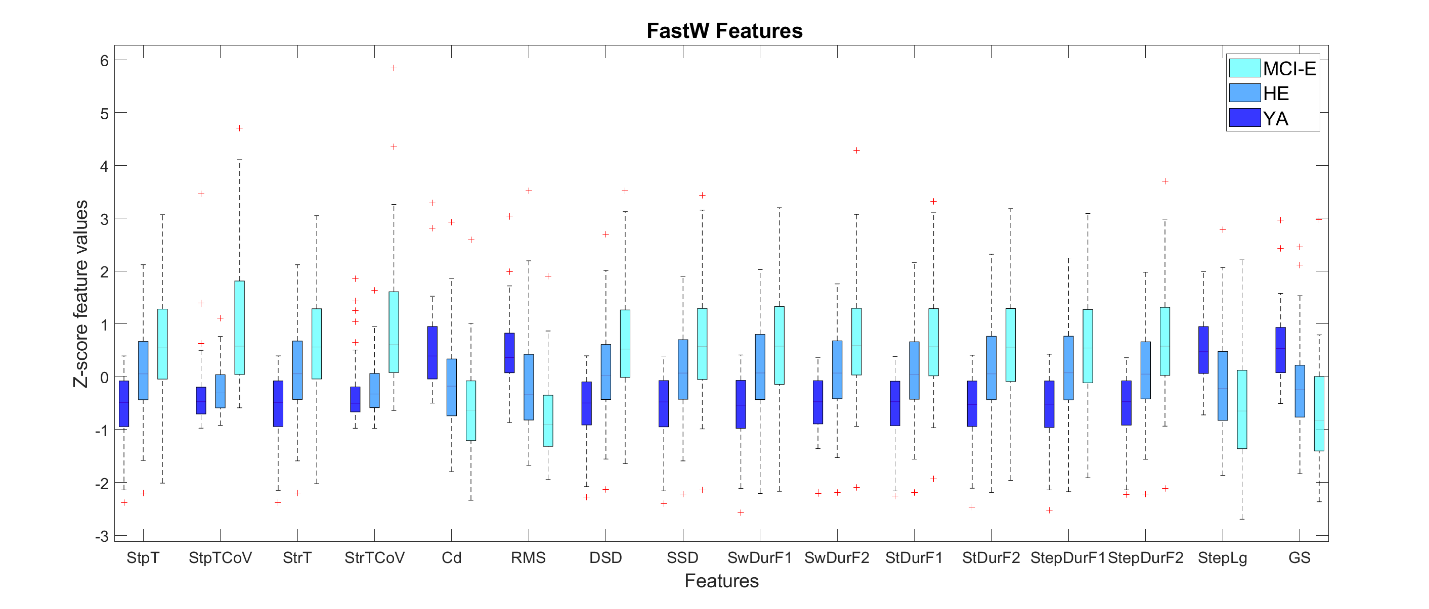

**Supplementary Figure 4.** Z-scores of STGF for the three groups in FastW walking task.

#### 1.2 Supplementary Tables

**Supplementary Table 1**. Results of the non-parametric permutation two-way analysis of variance test for each STGF, with GROUP and TASK as main effects, and the interaction between them.

|  | **F(2,488) group** | **p group** | **F (3,488) task** | **p task** | **F(6,488) interaction** | **p interaction** |
| --- | --- | --- | --- | --- | --- | --- |
| **StpT** | 60.60 | 0.00 | 67.00 | 0.00 | 1.78 | 0.17 |
| **StpTCoV** | 28.78 | 0.00 | 30.88 | 0.00 | 3.33 | 0.02 |
| **StrT** | 60.85 | 0.00 | 67.27 | 0.00 | 1.76 | 0.15 |
| **StrTCoV** | 29.87 | 0.00 | 29.53 | 0.00 | 2.53 | 0.05 |
| **Cd** | 61.58 | 0.00 | 91.70 | 0.00 | 0.83 | 0.47 |
| **RMS** | 50.90 | 0.00 | 84.75 | 0.00 | 2.27 | 0.07 |
| **DSD** | 26.83 | 0.00 | 42.32 | 0.00 | 2.22 | 0.08 |
| **SSD** | 63.90 | 0.00 | 62.35 | 0.00 | 1.67 | 0.16 |
| **SwDurF1** | 63.57 | 0.00 | 63.66 | 0.00 | 1.60 | 0.20 |
| **SwDurF2** | 61.64 | 0.00 | 58.77 | 0.00 | 1.72 | 0.15 |
| **StDurF1** | 52.42 | 0.00 | 61.55 | 0.00 | 1.92 | 0.13 |
| **StDurF2** | 54.22 | 0.00 | 65.73 | 0.00 | 1.81 | 0.13 |
| **StepDurF1** | 60.90 | 0.00 | 68.34 | 0.00 | 1.64 | 0.17 |
| **StepDurF2** | 57.78 | 0.00 | 62.98 | 0.00 | 1.82 | 0.15 |
| **StepLg** | 50.92 | 0.00 | 62.49 | 0.00 | 0.67 | 0.59 |
| **GS** | 63.55 | 0.00 | 99.00 | 0.00 | 1.12 | 0.33 |
